# Shared loci of telomere length with brain morphology, and pleiotropy in transcriptomic and epigenomic profiles of brain

**DOI:** 10.1101/2020.08.03.20167726

**Authors:** Gita A Pathak, Frank R Wendt, Daniel F Levey, Adam P Mecca, Christopher H van Dyck, Joel Gelernter, Renato Polimanti

## Abstract

Premature shortening of telomere length is observed in neuropsychiatric disorders. We tested genetic colocalization of seven and nine leukocyte telomere length (LTL) loci in two ancestry groups, European (EUR) and East-Asian (EAS), respectively (total n=60,601) with brain morphology measures for 101 region-of-interests (ROI) (n=21,821). The posterior probability (>90%) was observed for ‘fourth ventricle’, ‘gray matter’ and ‘cerebellar vermal lobules I-IV’ volumes. We found regulatory genes (p ≤2.47 x10^−6^) by integrating transcriptomic (EAS=4;EUR=5) and methylation data (EUR=17; EAS=4) of brain tissues using Summary-based Mendelian Randomization (SMR). The LTL SNP associations were tested for brain-based chromatin profiles using H-MAGMA (EUR=50; EAS=97; p ≤ 1.02 x10^−6^). Pathway enrichment of tissue-specific genes highlighted calcium ion transport (fetal brain) and G2/M cell cycle transition (adult brain). This study provides evidence that previously reported LTL associations with neuropsychiatric disorders could be related to a shared genetic relationship between LTL and brain structural and regulatory traits.

## 1 Introduction

Telomeres are short sequences of “TTAGGG” at the 3’ end of the chromosomes. With replicating cell cycles, telomere length continues to shorten and represents the mitotic history of the cell. The telomere length shortens (i.e., the number of TTAGGG repeat motifs decreases) over chronological age (i.e., actual age of the person) (Koliada et al., 2015). Additionally, under the effects of influences such as, inflammation, oxidative stress, elevated hormone levels under stress or pathological conditions, telomere length can shorten prematurely (Vakonaki et al., 2018). Perturbations in telomere lengthening enzymes such as telomerase reverse transcriptase (*TERT*) and telomerase RNA component result in cellular damage and genomic instability, further accelerating damage to physiological processes termed cellular aging (Bär and Blasco, 2016).

Psychiatric traits such as major depressive disorder (MDD), schizophrenia, anxiety, bipolar disorder (Muneer and Minhas, 2019), and neurodegenerative disorders (e.g., Alzheimer’s disease) have documented associations with shortened leukocyte telomere length (LTL)(Forero et al., 2016). Childhood anxiety and adversity contribute to lifetime diagnosis of mood disorders(McLaughlin et al., 2012) such as MDD(Kaczmarczyk et al., 2018; Wang et al., 2017), bipolar disorder(Aas et al., 2016), schizophrenia(Aas et al., 2019), and posttraumatic stress disorder(McLaughlin et al., 2017). Childhood adversities and chronic illness are correlated with shorter leukocyte telomere length in individuals with anxiety diagnosis (Kananen et al., 2010). In addition to brain-based disorders, LTL is associated with brain morphology of such structures as hippocampal, temporal, and cingulate region volumes among others (King et al., 2014). Teenagers with MDD were reported to have shorter telomeres and smaller right hippocampal volume (Henje Blom et al., 2015). A longitudinal study measuring the effects of various mental training modules (to improve cognition and behavioral traits) reported a significant association between reduced telomere length and thinning of the cortex structure highlighting neuronal plasticity variation between telomere and certain brain structures(Puhlmann et al., 2019). The association of telomere length variability with psychiatric disorders has led to the hypothesis of accelerated cellular or biological aging mediating physiological deterioration (Aas et al., 2019).

Telomerase mediates cell differentiation and is highly expressed in neural stem and progenitor cells, and declines with increasing age (Liu et al., 2018). Dysfunction in telomerase impairs neural circuitry in the hippocampus, which is one of many factors that leads to memory decline (Zhou et al., 2017). The expression of *TERT* is lower in astrocytes than oligodendrocytes (Szebeni et al., 2014), and telomere attrition induces cellular senescence of astrocytes leading to neurological dysregulation (Cohen and Torres, 2019). Telomere shortening disrupts the neuronal differentiation cell cycle (Ferrón et al., 2009).Neurogenesis in both adult and fetal tissue is influenced by functioning telomerase and interaction with sheltrin complex which protects the telomeric caps (i.e., prevents reduction in telomeric TTAGGG repeat number) (Lobanova et al., 2017). Even though telomere length is generally expected to be associated with age-related disorders, telomere length attrition can be influenced by maternal and intrauterine stressors during fetal development exhibiting its effects during an individual’s life course (Edlow et al., 2019).

Twin-based genetic heritability of LTL is estimated at around 30-60% (Dorajoo et al., 2019). Genome-wide association studies (GWAS) measure relative differences in frequencies of genome-wide single nucleotide polymorphisms (SNPs) for a trait of interest. A relatively high proportion of SNP associations are found in non-coding regions and may have tissue specific regulatory effects in local or distant regions (Giral et al., 2018). Tissue- and cell-type specific regulatory effects may be investigated by comparing the association of SNPs with gene expression (termed expression quantitative trait loci, eQTL) and methylation levels of CpG (cytosine-phosphate-guanosine) sites termed methylation QTL (mQTL) (T. Zhao et al., 2019). Leveraging large studies with genetic, transcriptomic, and epigenetic data allows us to identify putative tissue and cell-specific regulatory effects of trait-specific SNPs. To our knowledge, no study has investigated shared causal loci for brain morphology and brain tissue based multi-omic regulatory consequences of telomere length SNP associations.

In this study we used large scale LTL GWAS of 37,505 European and 23,096 Chinese individuals (n_total_=60,601) (Dorajoo et al., 2019) to identify a) LTL causal loci shared with brain volume measures, b) transcriptomic and epigenetic regulatory effects, and c) chromatin profiles in brain tissue (Figure 1). First we analyzed whether LTL risk loci are shared with 101 T1-MRI (magnetic resonance imaging weighted for longitudinal relaxation time) brain region-of-interest phenotypes (n=21,821) (B. Zhao et al., 2019).Then, we integrated meta-analyzed mQTL (Qi et al., 2018) (n = 1,160) and eQTL associations from different regions of the brain (n = 1,194) obtained from Genotype-Tissue Expression GTEx (GTEx Consortium et al., 2017), CommonMind Consortium (CMC) (Fromer et al., 2016), the Religious Orders Study and the Memory and Aging Project (ROSMAP) (Ng et al., 2017) and prefrontal cortex-based eQTL (n = 1,387) of PsychENCODE consortia (Gandal et al., 2018) were used to conduct gene based association study for LTL using Summary-based Mendelian Randomization (Zhu et al., 2016). Chromatin profiles from human brain tissue of two developmental stages – fetal and adult, and two cell types – astrocytes and neurons were investigated to identify genes specific to tissue and cell types using H-MAGMA (Hi-C-coupled multimarker analysis of genomic annotation) (Sey et al., 2020).

**Figure 1:**
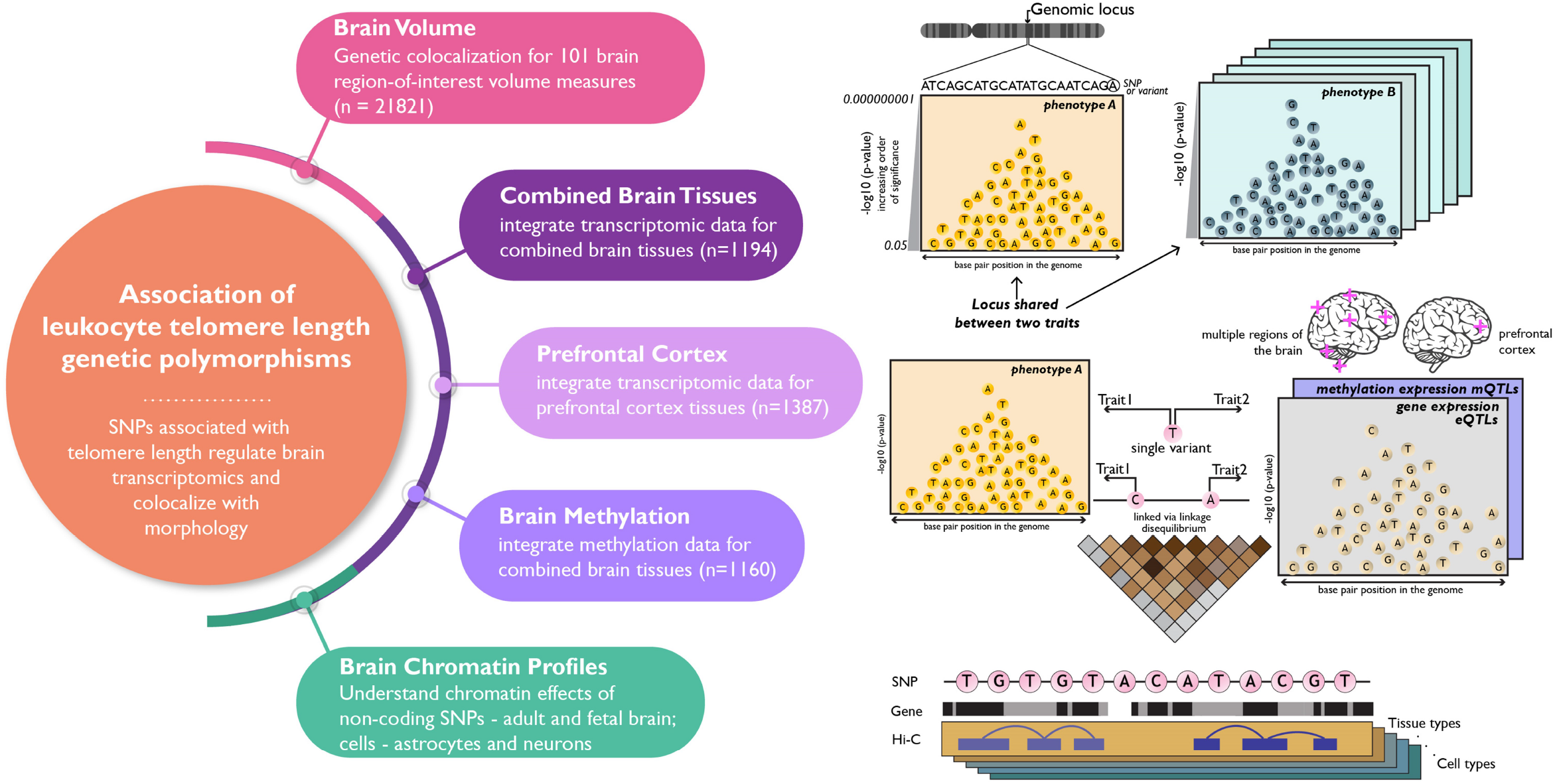
Study Overview: Schematic presenting the hypothesis and four major domains studied – i) shared genomic regions between leukocyte telomere length (LTL) and brain volume measures, and gene-based associations for LTL in brain tissue-based ii) transcriptomics – combined brain tissue (meta-analyzed from different regions of the brains) and prefrontal cortex, iii) methylomics – combined brain tissues (meta-analyzed), and iv) Hi-C chromatin profiles – adult and fetal brain, astrocytes and neurons.

## 2 Results

### 2.1 Genetic colocalization with brain morphology

We performed colocalization analysis using *coloc* (Giambartolomei et al., 2014) between LTL and 101 brain volume measures to identify causal genomic loci that with respect to telomere length and measures of brain morphology. The *coloc* method measures the posterior probability for five hypotheses (H_**0-**_H_**4**_), where H_**3**_ and H_**4**_ posterior probabilities (PP) represent sharing two and one genetic variant, respectively. For the EUR population, we observed four brain volumes with PP higher than 30%: left postcentral [PP_H3_= 1%, PP_H4_= 47% ; chr20(q13.33)], left caudal middle frontal [PP_H3_= 1%, PP_H4_= 51%; chr20(q13.33)], right precentral [PP_H3_= 73%, PP_H4_= 1%; chr2(p16.2)] and gray matter [PP_H3_= 99%, PP_H4_= 1% ; chr10(q24.33)]. The top variants associated with both traits in this locus [chr10(q24.33)] are rs11191849 and rs11191848, both mapping to*OBFC1* (alternatively known as *STN1*).

For the EAS population, we found six brain volume measures: gray matter [PP_H3_= 10%, PP_H4_= 32% ; chr11 (q22.3)]; right cerebellum white matter [PP_H3_=1%, PP_H4_ = 42% ; chr10 (q24 -q25.1)]; right basal forebrain [PP_H3_= 3%, PP_H4_ =39% ; chr20 (q13.33)]; cerebellar vermal lobules I-IV [PP_H3_= 42%, PP_H4_ =50%; chr7 (q31.33)]; right supramarginal - [PP_H3_= 14%, PP_H4_ =65% ; chr10 (q24 -q25.1)], and fourth ventricle [PP_H3_= 4%, PP_H4_ = 96% ; chr20 (q13.33)](Figure 2). The strongest PP was seen on chr20 (q13.33) for sharing risk variants between LTL and fourth ventricle which is located dorsal to the brain stem and anterior to the cerebellum, and drains the cerebrospinal fluid (CSF) into the spinal cord (Roesch and Tadi, 2020). The top variant in this locus rs7361098 is in the intronic region of the *RTEL1* (regulator of telomere elongation helicase 1; hg19 and 38) and upstream to the *RTEL1-TNFRSF6B*. On chr7 (q31.33), the cerebellar vermal lobules I-IV had high aggregated probability (92%) of shared genomic risk locus between LTL and volume of cerebellar vermal lobule. (Supplementary file 2). The top variants in this locus were rs34204767 in*GPR37* and rs6968500 in *C7orf77* (upstream). The vermal lobules are located in the cerebellum midline and affect behavioral and cognitive phenotypes (Yucel et al., 2013).

**Figure 2:**
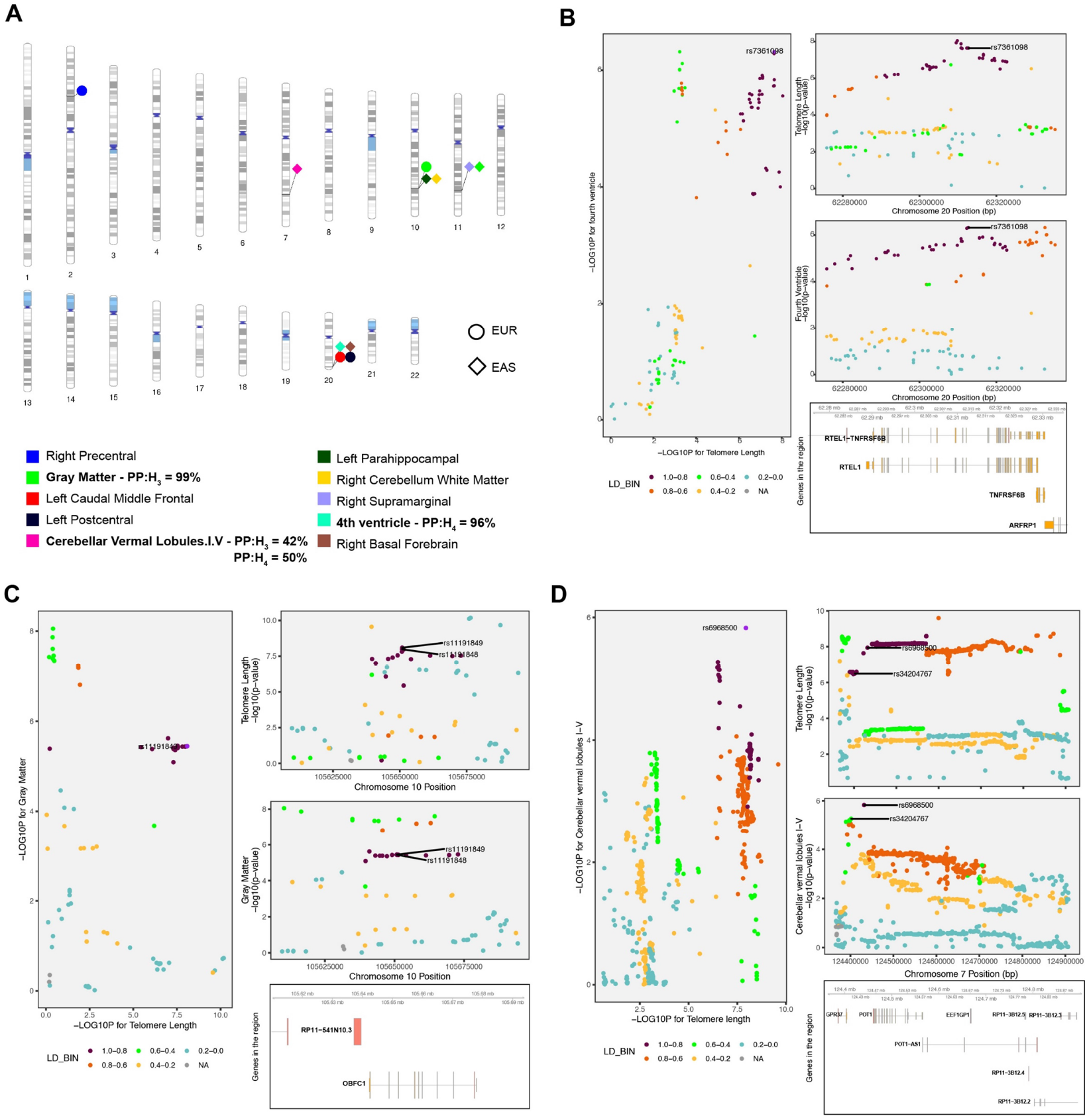
Genetic colocalization. **A)** The ideogram highlights genomic regions wherein genetic variants (SNPs) are shared between leukocyte telomere length (LTL) and respective brain volume measures whose PP >30%. Posterior probability >90% of sharing a causal variants for each LTL-brain volume measure pair is bolded. **B)** Regional association plot for LTL and fourth ventricle on chr20 (q13.33) and the top significant variants for both traits in the locus is shown followed by genes in the region (hg19 build). **C)** Regional association plot for LTL and gray matter on chr10 (q24.33) and the top significant variants for both traits in the locus is shown followed by genes in the region (hg19 build). **D)** Regional association plot for both traits on chr7 (q31.33) and the top two most significant variants in the locus are shown followed by genes in the region (hg19 build).

### 2.2 Brain tissue based transcriptomic and epigenetic QTLs

We integrated expression meta-analyzed cis-QTL data from different tissues of the brain (brain_**meta**_) with SNPs associated with LTL considering allele frequency and LD structure of the EUR and EAS population separately. For brain_**meta**_ eQTL, we found one bonferroni significant gene for the EAS population (*DHRS1*; p=1.55 x 10^−6^and p_HEIDI_ = 1.25 x 10^−21^). None of the genes were bonferroni significant for the EUR population. Significant p-value HEIDI test indicates a linkage model i.e. more than one variant that are probably in linkage, are associated with eQTL and telomere length, whereas non-significant HEIDI p-value indicate pleiotropy under single causal variant model (Figure 3A & 3B).

**Figure 3:**
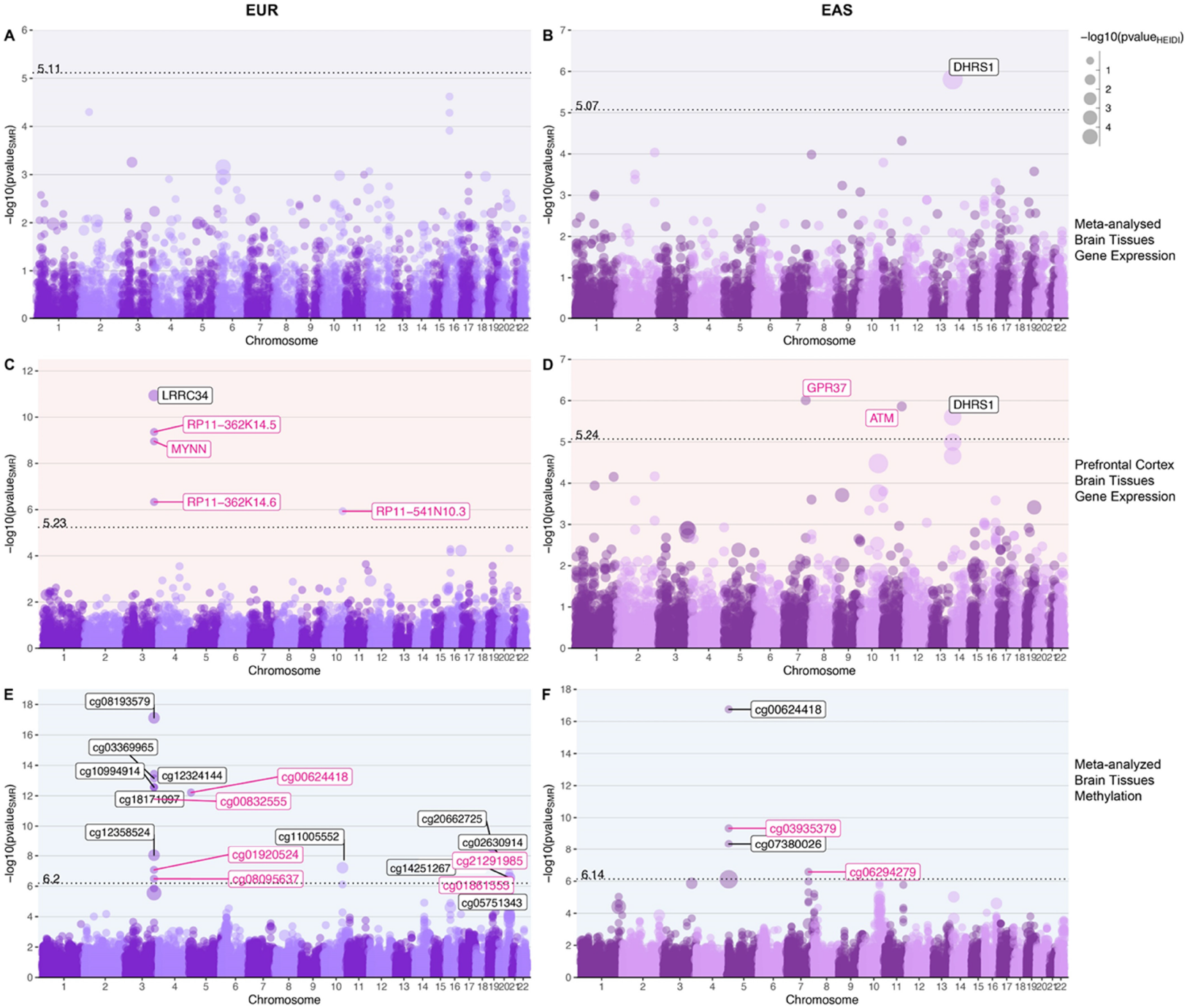
Gene-based associations integrating brain tissue eQTL and mQTL profiles with genetic variants associated with telomere length in EUR and EAS populations. A,B) Bokeh plots showing LTL-associated genes for gene expression (eQTLs) of combined brain tissues performed using SMR. C,D) Bokeh plots showing LTL-associated genes for expression in prefrontal cortex E,F) Bokeh plots showing LTL-associated genes for methylation in combined brain tissues. The left side panels show association in EUR population and EAS associations are shown on the right. The x-axis shows genomic coordinate for the gene/CpG sites and the y-axis shows the -log10 of p-value of SMR test, so the order of increasing significance moves vertically. The dotted line shows the Bonferroni significance and annotated with respective value on top of the line. The size of the dots are scaled to the -log10(p-value) of HEIDI test which tests for pleiotropy under single causal variant model defined as p-valueHEIDI >0.05 or -log10 (p-value) = 1.3 highlighted in pink text label.. Note: Bokeh plots are intersection between manhattan and bubble plots.

Integrating eQTL data from prefrontal cortex (eQTL_**pfc**_), we found five genes - *LRRC34* (p= 1.15×10^−11^;p_HEIDI_ = 9.64 x10^−4^), RP11-362K14.5 (p= 4.38 x10^−10^;p_HEIDI_ = 0.13), *MYNN* (p= 1.11 x10^−9^;p_HEIDI_ = 0.17), RP11-362K14.6 (p= 4.67 x10^−7^p_HEIDI_ = 0.17), and *RP11-541N10*.*3* (p= 1.18 x10^−6^; p_HEIDI_ = 0.10) in the EUR population. All the significant genes, except *LRRC34* had a non-significant HEIDI p-value, suggesting pleiotropic variants in the risk region sharing association with both eQTL_**pfc**_ and LTL. We found three genes associated in the EAS population; *GPR37* (p=9.75 x10^−7^; p_HEIDI_ =0.22), *ATM* (p= 1.38 x10^−6^; p_HEIDI_ =0.06) and *DHRS1* (p= 2.47 x10^−6^; p_HEIDI_ = 1.11 x10^−5^). Genes *GPR37* and *ATM* have non-significant HEIDI test statistics, suggesting pleiotropy(single-variant) in the region (Figure 3C & 3D) (Supplementary file 3).

For the brain tissue mQTL data integrated with LTL association statistics, we found 17 significant CpG sites (p ≤4.54 x10_^-7^_) in EUR ancestry subjects. Six of the 17 CpG sites were associated under the pleiotropic model - cg00624418 (p_HEIDI_ =0.82), cg00832555 (p_HEIDI_ =0.06), cg01920524 (p_HEIDI_ =0.27), cg21291985 (p_HEIDI_ =0.22), cg08095637 (p_HEIDI_=0.14)and cg01861555 (p_HEIDI_ =0.40). In the EAS population, we found four CpG sites; cg00624418 (p = 1.77 x10^−17^; p_HEIDI_ =0.01), cg03935379 (p= 4.60 x10^−10^; p_HEIDI_ =0.09), cg07380026 (p= 4.37×10^−9^; p_HEIDI_ =0.01) and cg06294279 (p= 2.65×10^−7^; p_HEIDI_ =0.35). Of these associations, cg03935379 (*TERT*) and cg06294279 (*GPR37)* are associated under the pleiotropic model (Figure 3E & 3F) (Supplementary file 3). Since methylation studies are mostly performed in blood, and methylation levels vary between tissues, we interpreted the CpG sites in blood and different brain tissues using BECon (Edgar et al., 2017) (Supplementary file 4 and Figure 4). Using the EWAS Atlas (Li et al., 2019), we investigated the eight CpG sites that showed pleiotropy, for their disease-based tissue variability of methylation levels (Supplementary file4).

**Figure 4:**
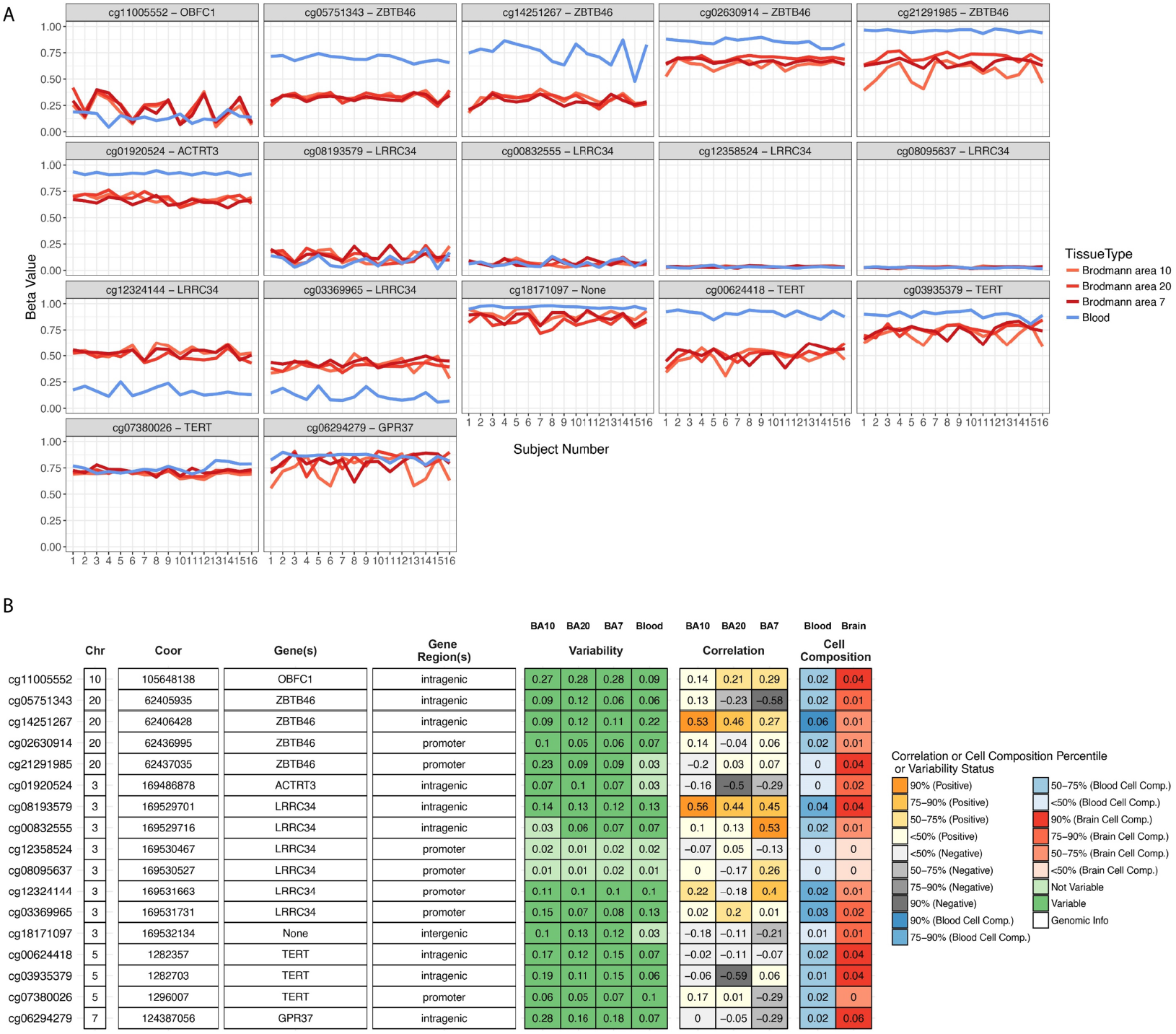
CpG site attributes and blood-brain tissue correlation. To aid in interpretation of the identified CpG sites, we used BECon for visualizing the inter-individual and tissue-based variability. **(Top)** The individual variability across three brain and blood tissue methylation levels is shown in each panel for respective CpG site from BECon data. **(Bottom)** The panel shows genomic location and annotation of the CpG sites and correlation value of blood methylation levels with each of the three brain tissues methylation. Tabular information is listed in Supplementary file 4.

### 2.3 Brain chromatin association for tissue and cell-types

We performed association analysis for LTL, by combining Hi-C chromatin SNP-gene associations for developing brain – fetal brain, dorsolateral prefrontal cortex – adult brain, and cell types – astrocytes and neurons using H-MAGMA (Sey et al., 2020). For fetal brain, we observed 21 genes (EUR) and 50 genes (EAS); adult brain (EUR-23; EAS-53); astrocytes (EUR-23; EAS – 53); neurons positive [cells sorted as NeuN+] (EUR – 21; EAS-54) and neurons negative [cells sorted as NeuN-] (EUR-25; EAS-55) (Figure 5).Several genes overlapped across tissue and cell types (unique number of genes - EUR= 50, significant genes lower than p 1.02 x10^−06^; EAS=97; significant genes lower than p 9.71 x10^−07^), and their specific distribution is shown in the Venn diagram (Figure 5) (Supplementary file5). Combining genes across both populations and considering those unique to each tissue and cell type, we found unique genes for fetal brain (eight genes), adult brain (nine genes), astrocytes (nine genes), neurons ; NeuN+ (11 genes) and NeuN-(15 genes) (Table 1). These genes were further investigated for enriched gene ontology pathways. We observed processes; calcium ion transport (p_FDR_ =0.012; fetal brain) and G2/M cell cycle transition (p_FDR_ = 0.036; adult brain) at p_FDR_<0.05 (Figure 5 top-right panel) (Supplementary file5 Table S6 and S7). To further the interpretation, we analyzed these genes using the whole-brain specific protein-protein interaction of all unique genes to tissue or cell types. The network was filtered to retain at least two-degrees, meaning that gene (node) should be connected to at least two other genes (nodes), to avoid selecting orphan/singular branches. This approach prioritizes genes that interact the most in a given network (Figure 5 bottom-right panel). This network revealed 12 intermediate genes that connected our query genes.

**Table 1:** List of genes unique or non-overlapping among the tissue and cell types. The genes highlighted in bold drive enrichment results discussed in the main text.

| Fetal Brain | Adult Brain | Astrocytes | Neurons (Both NeuN cells) |
| --- | --- | --- | --- |
| *RPL22L1, DCAF4, ZNF431*<br>*ZNF492, **CUL5, JPH4***<br>*TSSK4, DHRS1* | ***PRKCI, SKIL, NPY1R,***<br>***CEP72, CTC-457E21.9,***<br>***EPHX1, SEC62***<br>***PSME1, RP5-963E22.5*** | *RP11-678G14.2, ZNF98,*<br>*HAR1B, RP5-963E22.4,*<br>*HELZ2, SLC12A7, RP11-*<br>*807H17.1, AC005276.1,*<br>*BCL2L2* | *RP11-379K17.4, SRMS,*<br>*ACBD3, RP5-1087E8.2,*<br>*ZDHHC11, HYALP1*<br>*HYAL4,*<br>*ENSG00000269042,*<br>*ZFHX2,*<br>*ENSG00000274002, RP11-*<br>*358D14.2, GPR160, RP11-*<br>*225H22.4, PAPLN,*<br>*ENSG00000283755,*<br>*ZNF493, CTD-2561J22.5,*<br>*ADCK3, BRD9, WASL,*<br>*RP11-420H19.2, TPM4P1,*<br>*RP11-416N2.3, ALKBH8,*<br>*RP11-708B6.2, COL9A3* |

**Figure 5:**
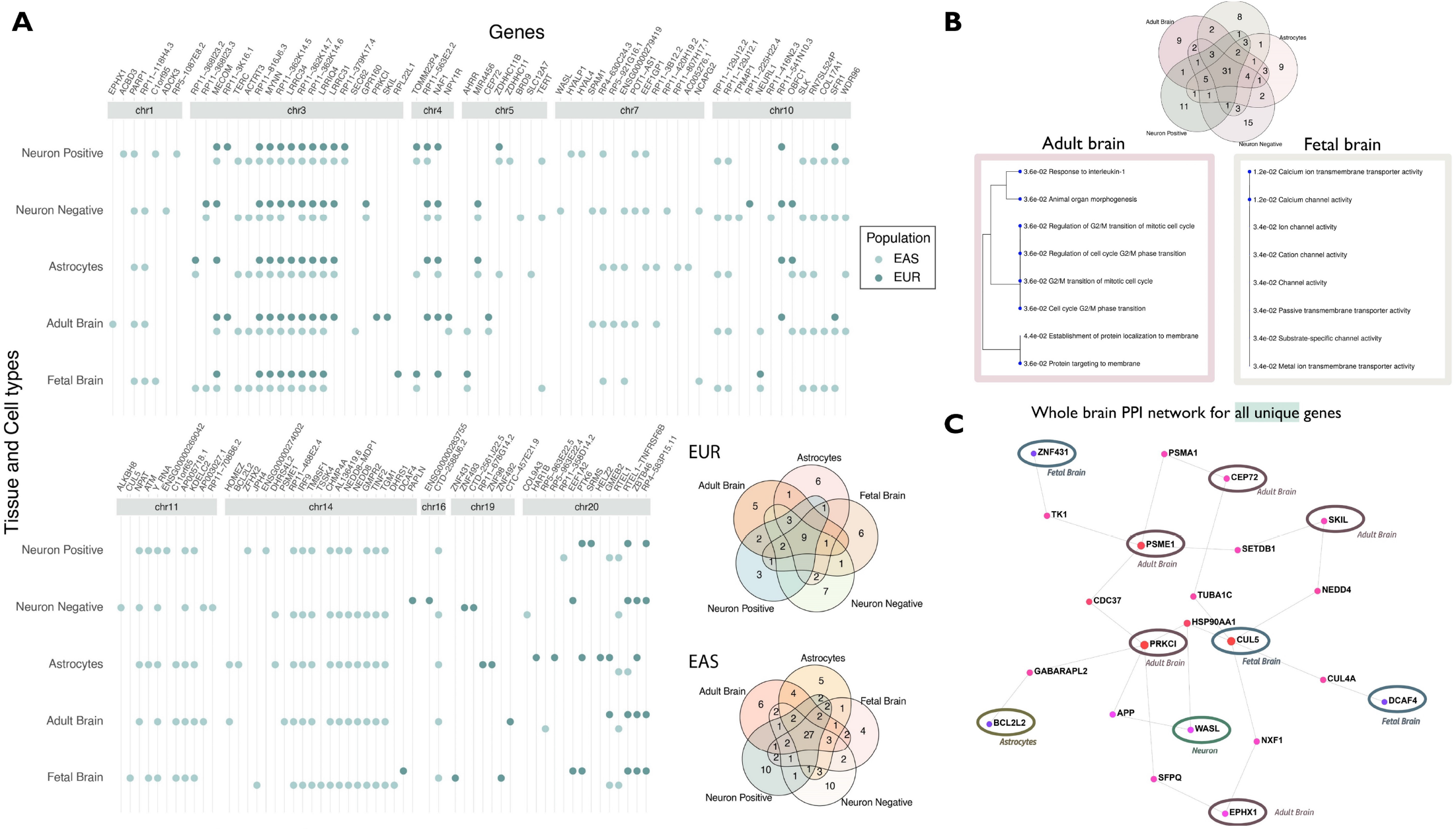
(A)Genes associated with LTL for tissue (fetal and adult brain) and cells (astrocytes and neurons). The genes are shown as circle data points categorized by tissue and cell types (x-axis) and arranged by genomic position (ascending position; y-axis). The dark green circles show significant genes for the EUR reference panel and EAS is shown as light green. The venn diagram on the right shows the distribution of distinct and overlapping genes for each population. Details are provided in Supplementary file 5. The Venn diagram on top right panel shows the distribution of combined significant genes in tissue and cell types. **(B) Gene enrichment analysis of distinct genes**. The genes that were unique to each tissue/cell type were analyzed for gene ontology enrichment. The processes are shown as dendograms with their pFDR value on the blue nodes. (C) **PPI network for all tissue/cell-type distinct genes**. The whole-brain specific PPI (protein-protein interaction) network for neuron genes is shown on the right, with query genes circled. Details of gene enrichment are reported in Supplementary file5 Tables S6 and S7.

### 2.4 Gene-drug interaction and enrichment of gene functions

We analyzed all significant genes identified from analyses for gene-drug interaction. There were 14 genes on whom the drug interaction was available (Table S1 in Supplementary file 6). To connect the influence of these drugs on gene functions weperformed drug set enrichment analysis. Out of 80 drugs, 10 drugs had associated information with gene function sets. There were 24 nominally significant (p<0.05) gene functions processes and one process i.e. transmission across chemical synapses was FDR significant (p= 0.00028; p_**FDR**_=0.019) (Fig S1 and Table S2 in Supplementary file 6).

## 3 Discussion

In this study, we investigated the role of genetic variants associated with leukocyte telomere length (LTL) in the context of brain-based expression, methylation, and chromatin profiles of two tissue types and three cell types. Analyzing different brain morphological traits, we discovered that the volumes of global gray matter, fourth ventricle and cerebellar vermal lobules I-IV, have genomic causal loci in common with LTL. The shared locus with global gray matter reported in our results corroborates previous findings of LTL association with cortical gray matter (King et al., 2014). A study of 15,892 individuals including subjects diagnosed with six psychiatric disorders showed gray matter loss across segmental brain regions in comparison to controls (Goodkind et al., 2015). The fourth ventricle bordered by the brainstem and the cerebellum, is one of the sites for CSF production and is the last ventricle connecting to the subarachnoid space (Roesch and Tadi, 2020). Pathologies involving the fourth ventricle are associated with abnormal CSF volume and flow (Whedon and Glassey, 2009). Different neuropsychiatric disorders such as schizophrenia have demonstrated enlarged fourth ventricle – which reflects a deficit in bordering tissue when compared to brain samples from healthy subjects (Juuhl-Langseth et al., 2012). Schizophrenia also has association with reduced telomere length (Russo et al., 2018). Furthermore, an extreme pathology (i.e. congenital deformity of the fourth ventricle) causes Dandy-Walker malformation which exhibits some symptoms with similarities to those seen in schizophrenia, obsessive compulsive disorder, and type-I and type-II bipolar disorder (Lingeswaran et al., 2009). Furthermore, the cerebellar vermal lobules I-IV are the top four anterior lobes connecting both hemispheres of the cerebellum, and are involved in motor, cognitive, and behavioral processing in autism spectrum disorders(D’Mello et al., 2015).Taken together, our results further corroborate the relationship between CNS morphology and telomere length.

The brain transcriptomic integrated association studies highlighted genes that are involved with both LTL and neuronal effects. *GPR37* (G-protein coupled receptor 37) is expressed in oligodendrocytes in different tissues of the brain contributing to myelination regulation (Smith et al., 2017). *GPR37* is associated with telomere length and serves as stress response neuroprotective gene. In major depression disorder, *GPR37* expression levels in brain tissue were lower compared to healthy participants (Mamdani et al., 2015). Furthermore, CpG site - cg06294279 located in the *GPR37* intragenic region was also associated under pleiotropic genomic risk (i.e. sharing association with both methylation level of the site and telomere length). The top variants in the colocalization analysis between LTL and cerebellar vermal lobules overlap with *GPR37*. The BECon databank shows that Broadman areas 7, 10, and 20 share little to low negative correlation with blood-based methylation in healthy individuals. To understand the role of the CpG site in diseases, we mined the EWAS Datahub (Li et al., 2019); CpG site - cg06294279 showed high variability in different brain tissues between cases and controls, for schizophrenia and Alzheimer’s disease (Supplementaryfile4).*LRRC34* (leucine rich repeat containing 34) gene expression is high in stem cells which lowers upon cell differentiation(Lührig et al., 2014). Individuals with Bipolar disorder who were on lithium showed changes in SNP-based transcriptome effect of *LRRC34* (Coutts et al., 2019). Furthermore, we found six CpG sites in the *LRRC34* gene that displayed mQTL associations for SNPs associated with LTL, out of which, site-cg08095637 had single-variant pleiotropic association.

Among genes identified from epigenomic integrated associations, *PARP1* (Poly(ADP-Ribose) Polymerase 1) is a regulator of DNA repair and cell death taking part in neurogenesis of neuronal and embryonic stem cells (Hong et al., 2019). Lowering the expression levels of *PARP1* is linked with abnormal ventricle volume in the brain and exhibits psychiatric symptoms including anxiety, depression, and behavioral impairment (Hong et al., 2019; Sriram et al., 2016). *ATM* (Ataxia Telangiectasia Mutated) is also involved in cellular DNA repair and differentiating pluripotent stem cells (Corti et al., 2019). ATM regulates GABAergic based neuronal plasticity and brain development and its dysregulation is associated with onset of different psychopathologies (Pizzamiglio et al., 2016). The mQTL analysis of brain tissue found CpG sites in addition to *LRRC34*, in the *TERT* gene. Methylation based biological age acceleration is associated with *TERT* and genetic overlap with schizophrenia, bipolar disorder, and dementia (Lu et al., 2018). *TERT* is also involved in paternally inherited epigenetic imprint observed in advanced paternal age which is a risk for several neuropsychiatric disorder (Hehar et al., 2017).Methylation levels of *ZBTB46* are associated with cognitive function (Starnawska et al., 2017) and schizophrenia in a RNA-sequencing gene expression study(Maycox et al., 2009; Zhang et al., 2020).

By integrating chromatin profiles for brain at two stages of development and different cell types, we found several overlapping genes across brain tissue and cell specific genes. For gene specific to Hi-C associations from fetal brain, we observed calcium ion transport process; this enrichment was driven by *JPH4*, and *CUL5. JPH4* belongs to the junctophilin family, is primarily expressed in neurons and regulates calcium signaling in normal brain development (Landstrom et al., 2014). *CUL5* (cullin5) encodes the vasopressin-activated calcium-mobilizing receptor (VACM-1) and is responsible for chromosomal stability and DNA repair mechanisms (Tapia-Laliena et al., 2019). Neural stem cells are precursors of neurons and glial cells in the brain and periodic responses from calcium signaling drive cell-specific neurogenesis (Toth et al., 2016). In the adultbrain, we observed enrichment of the G2/M cell cycle transition process. While neurons are generally not believed to enter M-phase, cell cycle dysregulation arising from pathological conditions can lead to undifferentiated neurons back in to the cell cycle and increase vulnerability to cell death (Walton et al., 2019). Reactivation of the cell cycle in adult neurons occurs in the circumstance of degeneration or injury to the central nervous system(Frade and Ovejero-Benito, 2015). Two genes driving cell cycle ontology were *EPHX1* and *PSME1*. Both *EPHX1* and *PSME1* are high expressed in the brain, *EPHX1* regulates endocannabinoid 2-arachidonoylglycerol in depression, stress, Alzheimer’s disease, and methamphetamine-dependence (Nithipatikom et al.,2014. Václavíková et al., 2015). Lower transcript levels of *PSME1* (proteasome activator subunit 1) have been observed in individuals with major depression disorder compared to healthy/control population (Redei et al., 2014).

We also observed several brain-based mQTLs which showed pleiotropic (single variant) association with LTL SNPs and several genes that were significant when integrating chromatin profiles. These results are in line with the genomic link between *TERT* and accelerated biological aging derived from methylation expression (Lu et al., 2018). The drug-gene interaction of significant genes showed enrichment for different transporter systems of chemical synapses, aquaporins and mitochondrial tRNA aminoacylation and transmission in the postsynaptic cell (Fig S1: Supplementary file 6). Together, these findings indicate the set of LTL-associated genes identified herein are associated with brain-related traits due to their regulatory effects on brain tissues and cells. Therefore, we believe telomere length genomics is an important avenue for future studies of mental health.

Most significant genes from one population are nominally significant in the other. However, there were few regions that are significant in one population, but not the other. These differences are due to recombination of the SNPs and allele frequency differences in the locus. Five loci on chromosomes 3, 4, 5, 10 and 20 overlap in both populations, however, the SNPs in these regions show differences in linkage disequilibrium highlighting the importance of studying the population based genetic association. These differences in SNP-peaks are visualized in genome-wide and regional plots (Supplementary file 1).

While our study has provided key evidence of shared genomic causal loci among telomere length, brain morphology, and regulatory traits, it has limitations. The results showing genetic co-localizing cannot be interpreted as mediating the direction of causality. Furthermore, colocalization of two traits under a single variant is a conservative model of testing, and there is biological plausibility of more than one variant exhibiting small effects. There are several SNPs in a locus exhibiting different effect directions for LTL and brain volume (Fig S2: Supplementary file 6). Therefore, it is difficult to assess the phenotypic consequences of each SNP because variants may alter physiological states in tissue or cell-type specific manner. We assessed regulatory changes using two populations and therefore the results may not apply to other major populations and will require separate investigation. Further studies using animal models would narrow targets with specific cellular changes. Our next steps will involve investigating the role of telomere length with respect to neuropsychiatric brain-disorders to deepen the understanding of cellular aging in neurological outcomes.

## 4 Conclusion

We identified shared genomic causal loci among those affecting telomere length, brain morphology and regulatory traits. The identified genes show involvement in the central nervous system, and provide evidence that shared genetics could explain, in part, the association of LTL with several brain-based outcomes reported previously. These results highlight that telomere length may serve as a marker for certain brain-based diseases such as psychiatric disorders. These findings additionally corroborate that in addition to methylation based biological aging, telomere-based cellular aging has the potential to provide further resolution on biological mechanisms of brain disorders.

## 5 Methods

All data used in this study were made publicly available by the cited consortia. This study was exempted from requiring institutional review board approval due to its use of de-identified aggregated data.

### 5.1 Cross-ancestry comparison of LTL-associations

We used the largest available LTL GWAS for EUR and EAS population. There are seven genome loci in EUR population, and nine independent loci in EAS population associated with leukocyte telomere length (LTL) (Dorajoo et al., 2019). Genomic risk regions for LTL were identified using FUMA (default values; LD -0.6 and 250kb locus) (Watanabe et al., 2017). A locus contains several SNPs independent significant SNPs by merging LD blocks within 250kb. Loci on chromosomes 2 and 19 that are observed in EURs are not present in EAS population. Similarly, there are loci specific to EAS population are on chromosomes 1, 7 and 14. Regions which are positionally similar in both populations are on chromosomes 3, 4, 5, 10 and 20. Length of these genomic regions varies by population due to LD structure and different number of significant variants within these regions. Therefore, in this study we investigated pleiotropic associations of SNPs with LTL and quantitative expression of brain tissues in both populations (Supplementary file1).

### 5.2 Brain morphology colocalization

To evaluate which loci are shared between LTL and brain volume measures, we used the largest available GWAS of 101 phenotypes of brain morphology(B. Zhao et al., 2019). In this study, the brain volume measures or region-of-interest (ROI) were quantified using magnetic resonance imaging and mapped to catalogued ROI to identify (i.e. map them to) structural measures (B. Zhao et al., 2019). The GWAS of 101 brain morphology/region-of-interest measures includes meta-analysis of five cohorts – the Human Connectome Project (HCP) study, the Pediatric Imaging, Neurocognition, and Genetics (PING) study, the Philadelphia Neurodevelopmental Cohort (PNC) study, and the Alzheimer’s Disease Neuroimaging Initiative (ADNI) study (n=21,821). The GWAS association statistics for each of the 101 brain morphology measures (B. Zhao et al., 2019) were evaluated with respect to GWAS of LTL (Dorajoo et al., 2019) to identify which causal risk loci were shared across both traits using *coloc* (Giambartolomei et al., 2014). We performed pairwise colocalization analysis for each of the risk locus between LTL and each brain volume phenotypes using the appropriate 1000 Genomes Phase 3 reference panels (i.e., EUR and EAS). The *coloc* method in R-v3.6, evaluates posterior probability (PP) for four alternative hypothesis – A) H_**1**_: association with trait 1, not with trait 2, B) H_**2**_: association with trait 2, not with trait 1, C) H_**3**_: association with trait 1 and trait 2, two independent SNPs, and D) H_**4**_: association with trait 1 and trait 2, one shared SNP (Giambartolomei et al., 2014). We used the coloc.abf function using p-values, respective ancestry’s allele frequencies and sample size with default priors. Here we report results for sharing single causal variant with both traits (LTL and brain morphology) having H_**4**_ PP higher than 30% in the main text, detailed results can be found in supplementary file 2.

### 5.3 Transcriptomic and epigenetic QTLs in brain tissues

The association of LTL was integrated with eQTL and mQTL information derived from Genotype-Tissue Expression (GTEx) (GTEx Consortium et al., 2017), CommonMind Consortium (CMC) (Fromer et al., 2016), Religious Orders Study and the Memory and Aging Project (ROSMAP) and PsychENCODE consortia (Gandal et al., 2018). Data were included from two transcriptomic studies: a) multiple brain tissue i.e. meta-analysis of gene expression from different regions of the brain (brain_**meta**_) and b) prefrontal cortex (brain_**pfc**_), and methylation association study using meta-analyzed methylation expression from multiple regions of the brain using Summary-based Mendelian Randomization (SMR) (Zhu et al., 2016). Bonferroni correction (0.05/number of associations) was applied to identify significant gene associations for each of the six QTL analysis. We performed heterogeneity in dependent instruments (HEIDI) association analysis to test if a single variant is associated with phenotype and QTL (single variant model of pleiotropy). Significant HEIDI associations (p< 0.05) depart from this assumption of single variant association while non-significant associations (p > 0.05) do not reject the null, indicating a linkage model (i.e. different SNPs in linkage) (Zhu et al., 2016). The analyses were performed in each ancestry group (i.e., EUR and EAS) separately. Two sets of files were created using LD and SNP allele frequencies of each population from the 1000 Genome Phase 3 reference panel. This resulted in different number of SNPs considered for each population. Misaligned allele direction between QTL and LTL-SNP dataset were removed. The CpG sites which showed pleiotropic evidence i.e. single causal variant between LTL and brain mQTL were interpreted using BECon for methylation level correlation between brain tissues and blood (Edgar et al., 2017) and extracted from EWAS DataHub for different brain disorders (Li et al., 2019).

### 5.4 Brain chromatin profiles using H-MAGMA

The gene-based association study for brain-derived chromatin profiles for fetal and adult brain regions and cells (i.e. astrocytes and neurons) was performed using H-MAGMA (Sey et al., 2020). This approach aggregates genetic variants to nearest genes derived from Hi-C (high-resolution chromatin conformation) data of fetal developing cortex, adult dorsolateral prefrontal cortex, and cell-types - iPSC derived astrocytes and cells sorted as NeuN+ and NeuN-from anterior cingulate cortex(Rajarajan et al., 2018). The H-MAGMA approach aids in providing functional/regulatory effects of non-coding SNPs, and is a complementary to transcriptome-wide association studies and the *coloc* approach used in this study (Sey et al., 2020). Due to telomeres/TERT’s role in the central nervous system and strong PP seen with brain volume measures, we tested all five tissue profiles with LTL GWAS association statistics. Each tissue/cell type association was performed for EUR and EAS population reference panels. Bonferroni correction (0.05/number of associations) was applied to identify gene associations for each the five tissue/cell type analyses. The genes that were unique to each tissue or cell type were analyzed for pathway enrichment using ShinyGO (Ge et al., 2019). Additionally, all unique genes were investigated for their role in whole-brain specific protein-protein interaction network using NetworkAnalyst3.0 (Zhou et al., 2019) retaining genes with at least two genes. The visualizations were created in ggplot, Gviz and PhenoGram (Wolfe et al., 2013). All significant genes from aforementioned analyses were investigated for gene-drug interactions using DGidb(Griffith et al., 2013). Then analyzed for gene function enrichment using DSEA(Napolitano et al., 2016).

## Data Availability

All the data used in this study are publicly available, hosted at the respective consortia portal. Chinese and European population telomere length GWAS (https://www.nature.com/articles/s41467-019-10443-2#data-availability); Brain Region Phenotypes GWAS (https://github.com/BIG-S2/GWAS);eQTL and mQTL data is available from SMR (https://cnsgenomics.com/software/smr/#Overview); H-MAGMA (https://github.com/thewonlab/H-MAGMA). The results generated in this study are provided in the main text and supplementary files

## 6 Acknowledgements

This study was supported by the National Institutes of Health (R21 DC018098, R21 DA047527, and F32 MH122058). We would like to acknowledge the participants and the investigators of all consortia’s whose data helped investigate the research question.

## 8 Data Availability

All the data used in this study are publicly available, hosted at the respective consortia portal. Chinese and European population telomere length GWAS (https://www.nature.com/articles/s41467-019-10443-2#data-availability);Brain Region Phenotypes GWAS (https://github.com/BIG-S2/GWAS);eQTL and mQTL data is available from SMR (https://cnsgenomics.com/software/smr/#Overview); H-MAGMA (https://github.com/thewonlab/H-MAGMA). The results generated in this study are provided in the main text and supplementary files.

## 11 Supplementary files

- **Supplementaryfile1**.**pdf:** Genome-wide and regional plots of LTL-GWAS in EUR and EAS population showing annotation of CADD scores, RegulomeDB, regulatory sites, and HiC chromatin interactions
- **Supplementaryfile2**.**xlsx:** Details of genetic colocalization between leukocyte telomere length (LTL) of EUR and EAS populations with 101 brain morphology measures
  - Table S1: Genomic locus in each population and corresponding five posterior probability values for each region reported in the manuscript
  - Table S2: Legend of brain volume measures analyzed with LTL
  - Table S3: Details of posterior probability values for all brain regions for Region1 (chr2:54461744-54498311) of EUR population
  - Table S4: Details of posterior probability values for all brain regions for Region2 (chr3:169392323-169599567) of EUR population
  - Table S5: Details of posterior probability values for all brain regions for Region3 (chr4:164001945-164126538) of EUR population
  - Table S6: Details of posterior probability values for all brain regions for Region4 (chr5:1279790-1355859) of EUR population
  - Table S7: Details of posterior probability values for all brain regions for Region5 (chr10:105608838-105694301) of EUR population
  - Table S8: Details of posterior probability values for all brain regions for Region6 (chr19:22089575-22319421) of EUR population
  - Table S9: Details of posterior probability values for all brain regions for Region7 (chr20:62421622-62467796) of EUR population
  - Table S10: Details of posterior probability values for all brain regions for Region1 (chr1:226376883-226653478) of EAS population
  - Table S11: Details of posterior probability values for all brain regions for Region2 (chr3:169360327-169582223) of EAS population
  - Table S12: Details of posterior probability values for all brain regions for Region3 (chr4:164007820-164126538) of EAS population
  - Table S13: Details of posterior probability values for all brain regions for Region4 (chr5:1276050-1343794) of EAS population
  - Table S14: Details of posterior probability values for all brain regions for Region5 (chr7:124366188-124907090) of EAS population
  - Table S15: Details of posterior probability values for all brain regions for Region6 (chr10:105671683-105801818) of EAS population
  - Table S16: Details of posterior probability values for all brain regions for Region7 (chr11:107983015-108367453) of EAS population
  - Table S17: Details of posterior probability values for all brain regions for Region8 (chr14:24623428-24730100) of EAS population
  - Table S18: Details of posterior probability values for all brain regions for Region9 (chr20:62275844-62335293) of EAS population
- **Supplementaryfile3**.**pdf**: Details of significant LTL associations for eQTL (gene expression) of meta-analyzed brain tissues and prefrontal cortex, and mQTL associations of meta-analyzed brain tissues in each population. Comparative results of all significant associations from one population are shown in other population as well.
- **Supplementaryfile4**.**pdf**: Visualization of mQTL sites that showed evidence of pleiotropy (single variant model from non-significant p-value HEIDI) were mined from EWAS DataHub in EWAS Atlas to show tissue-based variability in neuropsychiatric phenotypes
- **Supplementaryfile5**.**xlsx**: Details of chromatin-profiles associations for tissue and cell types in EUR and EAS population
  - Table S1: List of significant genes in fetal brain tissue for LTL associations in EUR and EAS population. Comparative results of all significant associations from one population are shown in other population as well.
  - Table S2: List of significant genes in adult brain tissue for LTL associations in EUR and EAS population. Comparative results of all significant associations from one population are shown in other population as well.
  - Table S3: List of significant genes in astrocyte cell types for LTL associations in EUR and EAS population. Comparative results of all significant associations from one population are shown in other population as well.
  - Table S4: List of significant genes in positive neurons cell types for LTL associations in EUR and EAS population. Comparative results of all significant associations from one population are shown in other population as well.
  - Table S5: List of significant genes in negative neuron cell type for LTL associations in EUR and EAS population. Comparative results of all significant associations from one population are shown in other population as well.
  - Table S6: List of genes that were distinct/non-overlapping in each tissue and cell-types and gene ontology enrichment results are shown next to the respective list of genes.
  - Table S7: Gene ontology enrichment results of the network from unique genes shown in Figure 6 (right panel).
  - Table S8: List of Bonferroni thresholds for each tissue and cell type in both populations and the number of significant associations.
- **Supplementary file 6**. Details of gene-drug interaction, drug set enrichment and effect direction of SNPs in the loci that are colocalized
  - Table S1: Gene-Drug interaction mined from The Drug-Gene interaction database
  - Fig S1. Drug Enrichment Set Analysis (DSEA). Analyzing enriched gene functions for drugs identified from drug-gene interaction (as shown above)
  - Table S2: Tabular associations of DSEA shown in the above figure
  - Fig S2: Effect direction of SNPs in the shared causal region for LTL and brain volume measures

